# A clinical decision support system for interventional urinary stone management planning

**DOI:** 10.1101/2023.06.28.23291929

**Authors:** Soorya Bhuphaalan, Sapthagirivasan Vivekanandhan

## Abstract

Urinary stone disease is a very common disease of the urinary tract where excessive mineral content in the kidneys form stones and obstruct the urinary tract, due to which affected patients experience severe pain and uneasiness. Identification of the appropriate interventional stone management method is usually done based on the size and location of the stone. Improper planning of the interventional method can lead to multiple revisits and unnecessary radiation exposure and thus an intelligent clinical decision system to precisely provide a treatment plan is required. In this paper, an intelligent system that recommends an appropriate interventional stone management system is proposed. Stone management data of 600 patients containing information about the stone size, location and the treatment provided to them was used to train machine learning models. The training and testing performance of different machine learning models with the dataset has been compared. Results showed that decision trees and support vector machines showed better results in predicting the right treatment planning method while given necessary inputs (stone size and location). This system can be useful in clinical setups in assisting urologists in planning treatment for urinary stone disease.

## 1. Introduction

Urinary stone disease (USD) or Urolithiasis refers to the obstruction of the urinary tract by mineral deposits, usually called urinary stones. USDs are a common disease characterised by heavy incruciating pain in the flank and abdomen region. USDs are commonly diagnosed by ultrasonography but other methods like intavenous pyelogram (IVP), non-contrast KUB-CT are also widely used [1]. Different procedures and guidelines have been followed for managing urinary stone disease. The common interventional methods in urinary stone management are extracorporeal shock wave lithotripsy (ESWL), percutaneous nephrolithotomy (PNL) and removal of stones through ureteroscopy (URS) [2]. Identifying the right urinary stone management method suitable for the patient and appropriate interventional planning is important to reduce radiation administered to the patient, improve recovery rate and avoid multiple revisits. The European Association of Urology (EAU) has provided guidelines for proper stone management by defining what procedures can be preferred for stones present in different regions of the urinary tract based on the size of the stone [3]. Similar guidelines have been defined by different associations [4]. In this paper, we describe an intelligent system based on machine learning to identify the suitable stone management technique for a patient from parameters like urinary stone size and location. This system can be used in clinical decision making for identifying the right stone management method from minimal inputs from the user.

## 2. Dataset

oThe dataset contains retrospective data of 600 patients who successfully underwent interventional stone management for urolithiasis at SRM Medical College Hospital and Research Centre, India. The data was collected with approval from the institutional ethics committee. Cases with more than one stone in the urinary tract were excluded from the dataset along with cases where interventional stone management procedures were repeated for the same patient. The stone management methods adopted for removal of urinary stones were in-line with the EAU guidance for interventional stone management. The dataset has 600 samples specifying details of the location and size of the stone and stone management technique performed for each patient. The summary of the dataset is given in table 1. Out of the 600 data samples 159 samples contained urinary stone in the top pole and middle regions of the kidneys, 160 samples contained urinary stone in the lower pole of the kidneys, 102 samples contained urinary stone in the proximal part of the ureter while 98 samples contained urinary stone in the distal part of the ureter. The remaining 81 samples contained stones in the urinary bladder region. The dataset was divided into 450 samples for training machine learning models and 150 samples for testing the performance of the trained models.

**Table 1.** Number of data samples in different location of the urinary tract.

| Location | Count |
| --- | --- |
| Upper pole and middle region of kidneys (Right and Left) | 159 |
| Lower pole of kidneys (Right and Left) | 160 |
| Proximal ureters (Right and Left) | 102 |
| Distal ureters (Right and Left) | 98 |
| Urinary bladder | 81 |

## 3. Methodology

Different machine learning algorithms are commonly being used as decision support tools in healthcare. In this study, performance of different machine learning algorithms for classifying urinary stones into different interventional stone management methods based on their size and location is studied. Four machine learning algorithms, k-nearest neighbours classifier, support vector machine classifier, decision tree classifier and random forest classifier were trained with the dataset, where location and size of the urinary stone as inputs were mapped to the stone management treatment provided as the target. Out of 600 samples 450 samples were used for training the machine learning models while the remaining 150 samples were used for validation. To assess the performance of the machine learning model, four metrics – accuracy, F1-score, precision and recall were calculated for the training and validation datasets. The training and validation performance metrics are shown in section 4.

### 3.1. k-nearest neighbours (kNN)

In kNN method, the distance between the input vectors in the feature space is used to classify inputs into different classes [5]. For each data sample (i), k-number of nearest data samples and their classification labels are obtained. The class label of the data sample is assigned as the labels that are the majority in the k nearest samples, i.e., mode of the class label of the k-nearest neighbors. The classification is complete when the class labels for all the data samples are calculated.

### 3.2. Support vector machine (SVM)

SVMs are popular supervised machine learning models used for classification [6]. Instead of comparing a large amount of data samples and calculating the distances between them, like in kNN method, SVM uses a line or boundary called decision boundary which is adjusted such that it divides the data samples into n classes. The decision boundary is an n-1 dimensional hyper-plane. SVM training is done by identifying the support vectors (data samples of a class closest to the data samples of another class) and defining a support plane passing through the support vectors. The distance between an initialized decision boundary and the support plane is maximized iteratively until all the support planes are at maximum distance away from the decision boundary. The final position of the decision boundary is fixed in the feature space and test data samples are classified by their position in feature space with respect to the decision boundary.

### 3.3. Decision trees

Decision tree classification is a commonly used supervised machine learning technique where input parameters are mapped to a defined class in a tree-like structure [7]. The input parameters are split at locations in the tree called nodes which branch into two or more paths that are connected to other nodes or an end output node called leaf node. After training, a decision tree is generated with nodes, branches and leaf nodes that can be used to test unknown data where the input data in analysed at different nodes in the tree until it reaches the final leaf node. Model homogeneity is achieved by minimizing the Gini Impurity Score, which is a measurement of the probability of a classification being false.

### 3.4. Random forests

Random forests classification involves the use of multiple decision trees to solve a classification problem. A random forest contains an ensemble of individual decision trees that are trained to classify an input into different classes. The input for each decision tree in the forest is a random subset of the input given to the random forest algorithm [8]. After each decision tree in the forest gives a classification output, the class that is returned as the output by the most number of decision is considered as the output class of the random forest algorithm.

## 4. Results

Different supervised machine learning models explained in the section 3 were trained and validated using the dataset defined in section 2. The machine learning models were built and trained using Python and OpenCV’s machine learning module. The results from the comparison of different machine learning model performance on the defined dataset is tabulated in table 2 and table 3.

**Table 2.** Training metrics for different machine learning methods.

| Method | Accuracy | F1-score | Precision | Recall | AUC |
| --- | --- | --- | --- | --- | --- |
| kNN (k=5) | 0.995 | 0.995 | 0.996 | 0.994 | 0.996 |
| Decision tree | 1.000 | 1.000 | 1.000 | 1.000 | 1.000 |
| Random forest | 1.000 | 1.000 | 1.000 | 1.000 | 1.000 |
| SVM (gamma =1.2) | 1.000 | 1.000 | 1.000 | 1.000 | 1.000 |

**Table 3.** Validation metrics for different machine learning methods.

| Method | Accuracy | F1-score | Precision | Recall | AUC |
| --- | --- | --- | --- | --- | --- |
| kNN (k=5) | 0.986 | 0.986 | 0.986 | 0.986 | 0.986 |
| Decision tree | 1.000 | 1.000 | 1.000 | 1.000 | 1.000 |
| Random forest | 0.990 | 0.995 | 0.990 | 1.000 | 0.996 |
| SVM (gamma = 1.2) | 0.993 | 0.996 | 1.000 | 0.993 | 0.997 |

The k-nearest neighbour model achieved a validation accuracy of 0.97 with k=5 and 0.93 with k=9. The model repeatedly achieved the highest accuracy with k=5. Decision tree model and random forests achieved accuracy of 1.000 on both training and validation datasets while the SVM model achieved a validation accuracy of 0.973 using radial basis function and a gamma value of 1.2.

The training and validation accuracy was calculated by obtaining the ratio of total number of correct predictions to the total number of predictions, as show in equation 1.

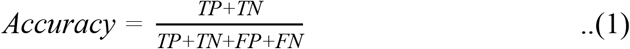

Other metrics such as recall, precision, F1-score and Area under the ROC curve (AUC) were also calculated for the training and validation data. The metrics were calculated as shown in equations (2), (3), and (4).

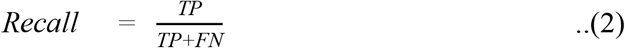

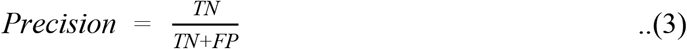

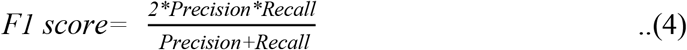

From the results it can be seen that the random forests, support vector and decision tree classification methods performed well compared to the kNN method. Decision trees method achieved the best performance without any overfitting.

## 5. Conclusion

Decision support systems in clinical setups have always been helpful in improving patient outcomes and reducing errors. A clinical decision support system for planning interventional stone management can help provide personalized care, improve stone free rate, reduce patient readmission rates, reduce unnecessary radiation exposure and unnecessary use of invasive procedures on the patient. Figure 1 explains a urinary stone treatment planning support system where abdominal CT or x-ray images are fed as the input from which stone size and location are calculated automatically or manually and fed into our subsystem where an appropriate stone management method is suggested according to the EAU guidelines for interventional stone management. From the results, it can be seen that decision trees and support vector machines showed the highest precision (100%) while the decision tree model performed the best among all other models. The machine learning models studied in this paper show promising results in accurately classifying the inputs (stone size and location) to identify the appropriate treatment method for managing urinary stones.

**Figure 1.**
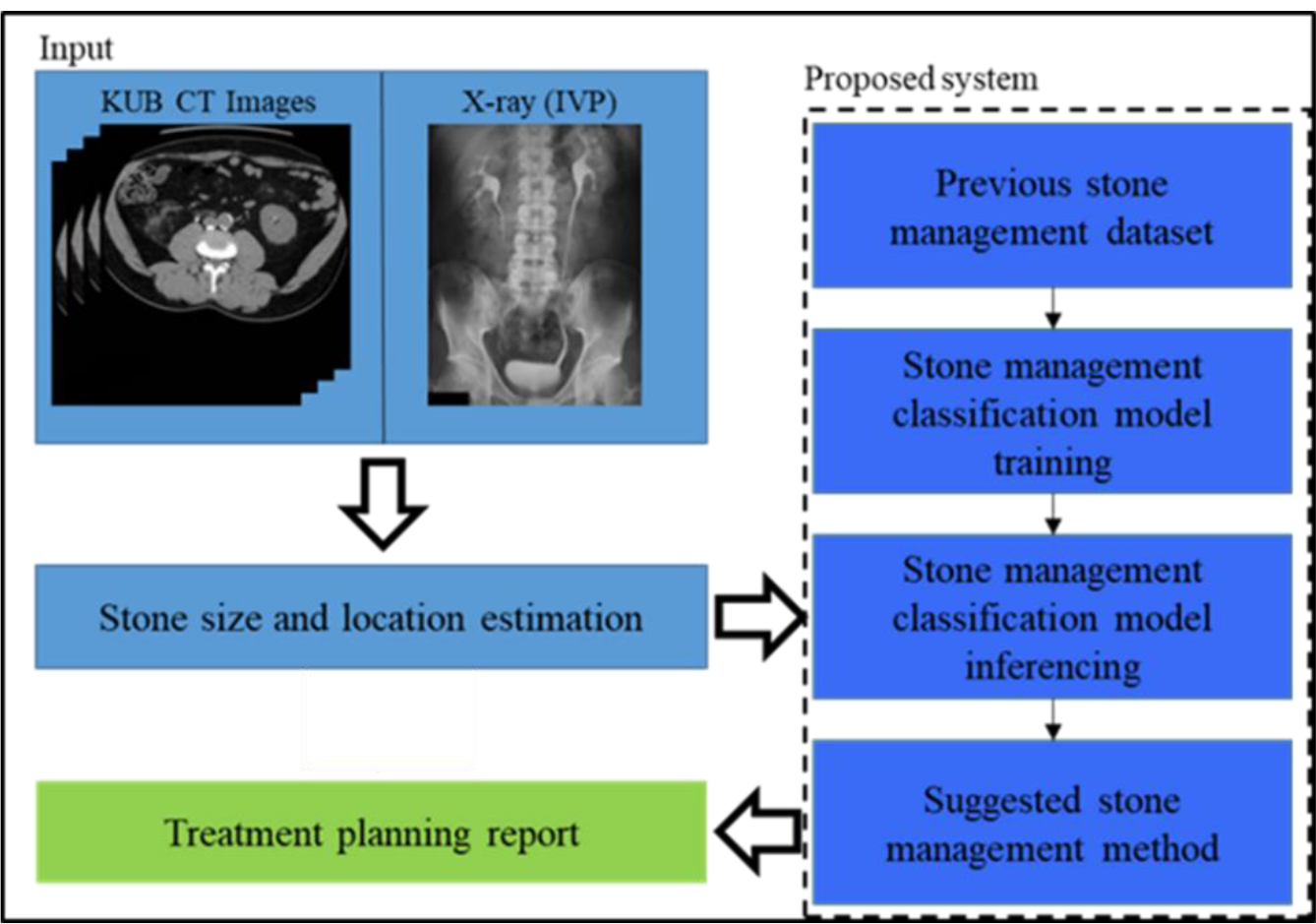
Proposed system as a part of a clinical decision making engine.

## Data Availability

All data produced in the present study are available upon reasonable request to the authors

